## Supplementary Information for "Cross-talk between red blood cells and plasma influences blood flow and omics phenotypes in severe COVID-19"

[1]Dynamics of Fluids, Department of Experimental Physics, Saarland University, Saarbrücken, Germany
[2]Institute for Clinical and Experimental Surgery, Campus University Hospital, Saarland University, Homburg, Germany
[3]Cysmic GmbH, Saarbruecken, Germany
[4]Department of Biochemistry and Molecular Genetics, University of Colorado Denver, Anschutz Medical Campus, Aurora, CO
[5]Department of Anesthesiology, Intensive Care, Emergency and Pain Medicine, University Hospital Wuerzburg, Wuerzburg, Germany
[6]Department of Anesthesiology, Intensive Care Medicine and Pain Therapy, University Hospital Frankfurt, Frankfurt, Germany
[7]Fraunhofer Institute for Translational Medicine and Pharmacology ITMP, Frankfurt, Germany
[8]Department of Physics and Materials Science, University of Luxembourg, Luxembourg City, Luxembourg
[9]Theoretical Medicine and Biosciences, Campus University Hospital, Saarland University, Homburg, Germany
[a]These authors contributed equally to this work.
[*]

### 1 Patient information

Supplementary Table S1 shows patient information, among which blood parameters measured the day of blood drawing for experiments.

### 2 Microfluidic flow analysis

At a given mean velocity, the distribution of the absolute value of the cell lateral position in y-direction, normalized by the channel width, is determined and the corresponding probability density function (pdf) is calculated. At low velocities, RBCs preferentially form axisymmetric croissants that flow in the channel center, while slipper-shaped RBCs that emerge at higher velocities flow at an off-centered equilibrium position (Figure S1A). We quantify the difference between pathological and healthy RBC flow by the so-called y-deviation, which relates the pdfs of the y-distributions of a given sample to the average distribution of healthy controls at specific cell velocities (Figure S1B).

### 3 RBC shapes in stasis

RBCs are also imaged in stasis for the assessment of shapes and cluster formation. Representative images from one healthy control donor and one COVID-19 patient in autologous and allogeneic plasma are given in Figure S2A. It appears clear that RBCs reverse their shape upon plasma exchanges, showing biconcave disks in control plasma (CinC and PinC) and sphero-echinocytes in patient plasma (PinP and CinP). Biconcave RBCs form organized rouleaux, in contrast with the sphero-echinocytes that are either separated or form disorganized clusters made of touching cells. The ratio of RBCs in clusters over the total number of RBCs quantifies the differences between CinC and PinP and shows the recovery of patient RBCs in PinC (Figure S2B, left panel). The number of RBCs per cluster in CinC is about twice the number of PinP. Notably, patient cell shape and rouleaux formation recovery in control plasma (PinC) results in no significant differences with control RBCs (CinC) (Figure S2B, right panel).

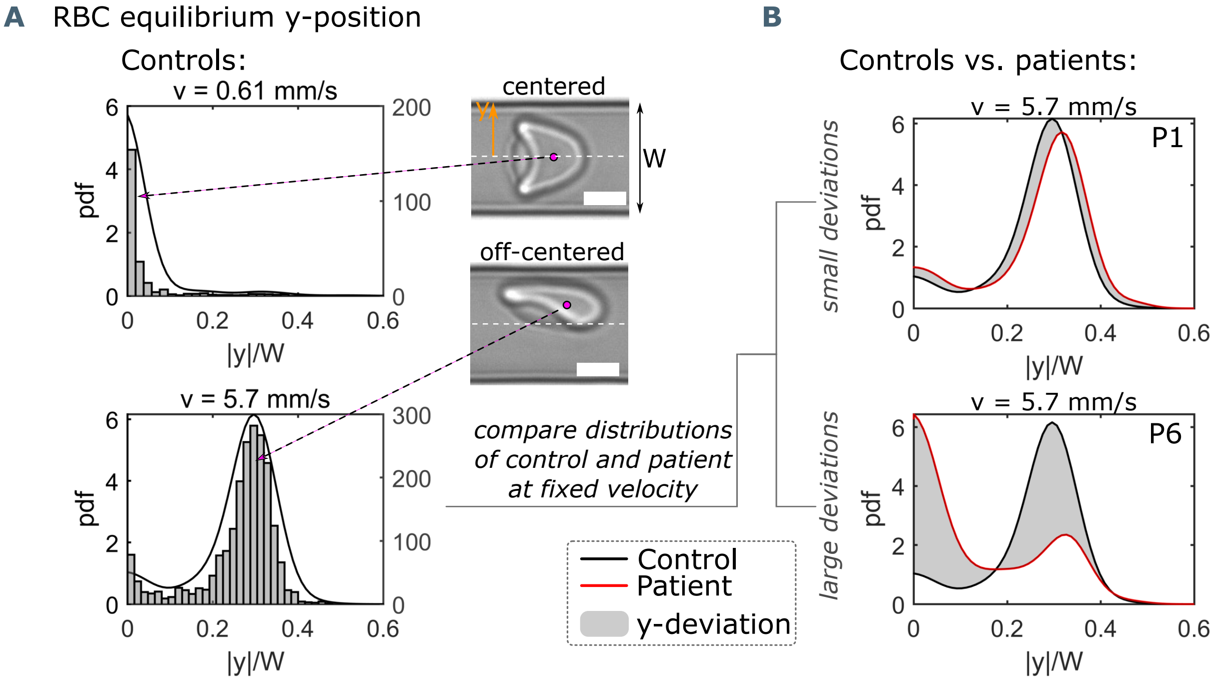


**Figure S1.** Single-cell flow characteristics. (A) Histograms and probability density distributions (pdfs) of the absolute value of the RBC y-position, normalized by the channel width W, at a representative low and high mean RBC velocity of 0.61 mm $s^{-1}$ and 5.7 mm $s^{-1}$, respectively. Scale bars represent a length of 5 µm. (B) Pdfs of the normalized y-position at 5.7 mm $s^{-1}$ for a healthy control and two patients (P1 and P6), indicating a small and large y-deviation (gray area) between the control and the corresponding patient.

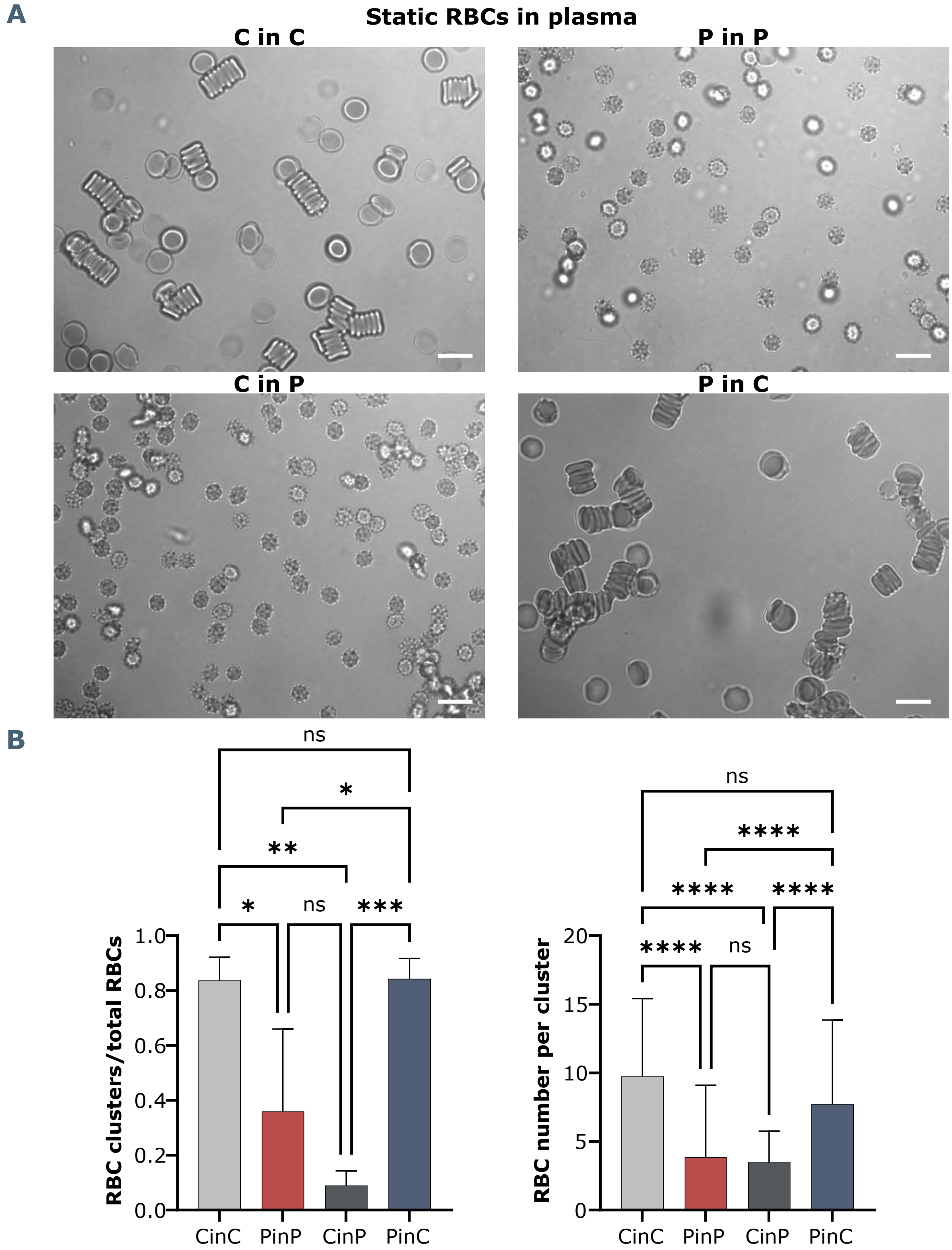


**Figure S2.** Brightfield microscopy and quantification of RBC shapes and clusters in stasis. (A) Representative RBC images from one healthy control in autologous plasma (CinC), one COVID-19 patient (PinP) and respective plasma exchanges, with control RBCs in patient plasma (CinP) and patient RBCs in control plasma (PinC). Scale bars represent 10 µm. (B) Quantification and statistical analysis by ANOVA from seven COVID-19 patients and three healhty controls. Left panel: ratio of RBC clusters over the total number of RBCs, right panel: number of RBCs per cluster.

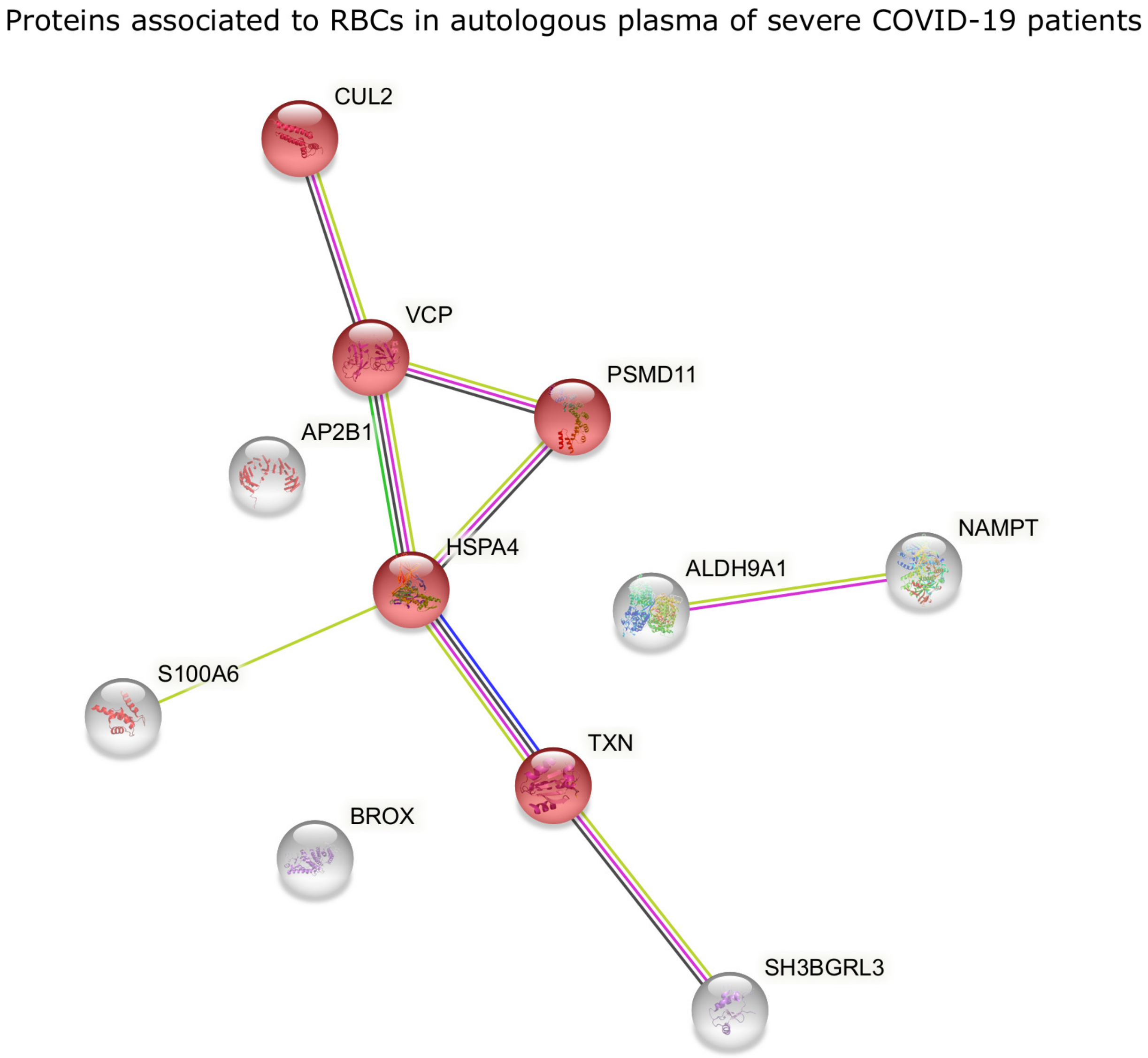


**Figure S3.** STRING analysis of proteins with significantly higher levels in RBCs of COVID-19 patients in autologous plasma. Evidenced in red are proteins associated with cellular responses to stress, involving ubiquitin-protein ligase complexes (CUL2), proteasome (PSMD11) and redox activity (TXN).

### Positively correlated proteins with sphero-echinocyte %

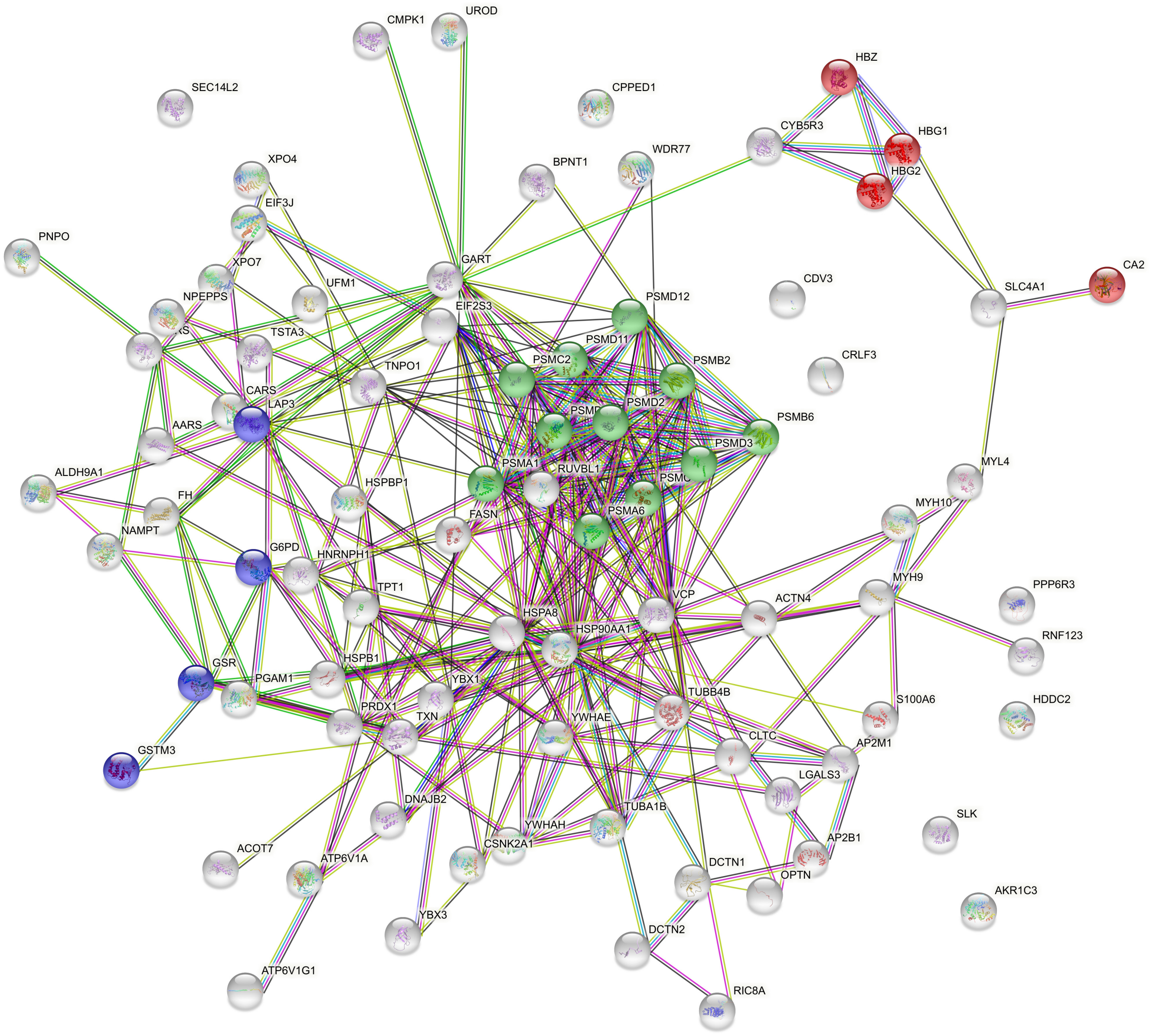


**Figure S4.** STRING analysis of positively correlated proteins with sphero-echinocyte percentage, highlighting the main processes involving pathological RBCs in COVID-19. Red: proteins involved in gas transport; green: proteasome-related proteins; purple: glutathione metabolism-related proteins.
